# The Impact of New SARS-CoV-2 Variants on Vaccine Breakthrough: A Pilot Study on Spreading Infection in the Communities

**DOI:** 10.1101/2021.09.21.21263898

**Authors:** Mohamad Ammar Ayass, Jin Zhang, Kevin Zhu, Wanying Cao, Natalya Griko, Victor Pashkov, Jun Dai, Trivendra Tripathi, Lina Abi Mosleh

**Affiliations:** Ayass Bioscience LLC, Frisco, Texas

## Abstract

**Background:** Coronavirus disease 2019 (COVID-19) vaccines are effective at helping protect against severe disease and death from variants; however, incident of breakthrough infection in vaccinated patients has been increased. Therefore, we aimed to assess the incidence of severe acute respiratory syndrome coronavirus 2 (SARS-CoV-2) new variants of concern in the communities and investigate vaccine breakthrough cases on our laboratory (Ayass Bioscience LLC) confirmed detection of COVID-19 variants in Dallas-Fort Worth (DFW), Texas.

**Methods:** Epidemiologic study has been performed at our laboratory. We studied the viral whole-genome sequence and genotyping analysis on 166 symptomatic cases of COVID-19 which were randomly selected from nasal swab positive cases assessed from June 1st to August 30th, 2021, by reverse transcription polymerase chain reaction (RT-PCR) cycle threshold (CT) values. COVID-19 variants were identified to be dominated by B.1.617.2 (89.2%) and followed by AY.3 (1.8%), B.1.1.7 (4.8%), a combination of B.1.526.1 and B.1.617.2 (3%), B.1.621 (0.6%), and P.2 (0.6%).

**Result:** The CT values showed significant difference among the three age groups: <30 years, 31-60 years, and >60 years by one-way ANOVA (N1: F (2, 118) =4.96, p=0.009; N2: F (2, 118) =4.95, p=0.009). No significant difference was observed by symptom, status of immunization, or vaccine manufacturer. A two-way ANOVA was performed to examine the effect of gender and variant group (Delta and other variants) on the CT values. The analyses revealed a statistically significant interaction between the effect of gender and variant group (N1, F (1.117) = 3.906, p = 0.05; N2, F (1, 117) = 7.402, p = 0.008).

**Conclusion:** Our study shows that Delta, the dominant variant of COVID-19, is spreading in the communities, and vaccine breakthrough cases occurred in the majority of Delta variant (91%) followed by AY.3 (5%), B.1.1.7 (2%) and 2% of the double variant of B.1.526.1 and B.1.617.2. The incidence of the breakthrough cases was not linked to a specific manufacturer. The CT value is likely to associate with age. This study also supports our laboratory’s ongoing efforts to sequence the SARS-CoV-2 virus from positive patient samples to identify the new viral variants and possible vaccine breakthrough mutations in the community.

## Introduction

Since the coronavirus disease 19 (COVID-19) pandemic started in March 2020, the severe acute respiratory syndrome coronavirus (SARS-CoV-2) virus has been mutating over time. New variants of the virus keep emerging occurring and circulating in the United State (CDC, variant proportion, 2021). There is no doubt that the new variants will bring the public health challenges and vaccine resistance concerns (Riemersma et al., 2021).

Recently, the Delta variant, also known as B.1.617.2, a viral lineage first identified in India was seen as the “ greatest threat” in the world’s effort to contain COVID-19 (Bolze et al., 2021). Delta variant has spread more easily and quickly than other variants which can lead to an increase in the number of cases, greater hospitalization, potentially more deaths, and reduce the effectiveness of vaccines (Riemersma et al., 2021, Brown et al., 2021, Chia et al., 2021).

The Food and Drug Administration (FDA) has approved Pfizer and Moderna booster shot to help against Delta variant in certain subpopulation and plan to introduce it to in general population later (FDA, 2021). However, would the booster shot work against a new variant such as Delta plus, Eta, Lota, Lambda, Mu? Currently the answer is not available. Thus, keeping track the variants and their trend is important to help us monitor and prepare for the potential surge in the communities. As a local laboratory, Ayass Bioscience has analyzed thousands of cases for COVID-19 and reported confirmed cases to the respective county health departments to help them respond to the COVID-19 pandemic. The purpose of this study is to assess the incidence of the COVID-19 variants in the communities and observe breakthrough vaccine cases.

## Materials and Methods

Specimens included in this study were collected from patients. There were 166 cases randomly selected from COVID-19 positive cases of real-time polymerase-chain-reaction (RT-PCR) assay run on the fully automated STEPONEPLUS System from Applied Biosystem. Nasopharyngeal swab specimens tested from June 1^st^ to August 30^th^, 2021. All testing and interpretation were conducted in accordance with the Patient Under Investigation (PUI) for 2019 novel coronavirus (2019nCoV). Two nucleocapsid genes of this virus (N1, N2) were detected in all cases. Human RNase P (RP) gene control values suggested sampling of patients and RNA isolation by Biomek I5 automated workstation (RNAdvance Viral Kit) were performed optimally.

The viral whole-genome sequence analysis was performed on all cases to identify SARS-Cov-2 variants by Genexus™ Integrated Sequencer system using Ion AmpliSeq™ SARS-CoV-2 Research Panel. Qualitative detection and differentiation of SARS-CoV-2 variants of interest was performed on Agena MassARRAY System using the MassARRAY SARS-CoV-2 Variant Panel v3 (RUO) protocol.

Meanwhile, a follow-up telephone call was performed as per as oral consent form guidelines of Ayass Bioscience Laboratory to address the questions of COVID-19 immunization, travel, and positive contact history, etc. 121 of 166 cases responded to our call and completed the questionnaires.

Data were summarized as the mean ± SD. Groups were compared by using one-way analysis of variance (ANOVA) and two-way ANOVA by the Turkey’s post-hoc test for multiple comparison. P-value ≤ 0.05 was considered to be statistically significant. Fisher’s exact test was also used. Analyses were performed on SPSS for Windows (version 23 Inc., Chicago, IL, USA).

## Result

### Patient characteristics

Of 166 patients, 47% of patients were male and 53% were female. The median age was 33 years (range from 4 months to 79 years). The age group was distributed as 0-17 years (24.1%), 18-29 years (19.9%), 30-39 years (23.5%), 40-49 years (15.7%), 50-59 years (7.8%), 60-69 years (6%), and 70 years (3%). The average RT-PCR cycle threshold (CT) value was 19.87±4.93 (N1) and 21.30±5.58 (N2), respectively. The more detailed information is listed below (Table 1). The viral whole-genome sequence analysis on 166 cases identified that COVID-19 variants were dominated by B.1.617.2 (89.2%) and followed by AY.3 (1.8%), B.1.1.7 (4.8%), double variant of B.1.526.1 and B.1.617.2 (3%), B.1.621(0.6%), and P.2 (0.6%) in the communities (Figure 1).

**Table 1:** 166 patients characteristics

|  | Delta variant | Other variants |
| --- | --- | --- |
| Gender: |  |  |
| Female | 80 (91%) | 8 (9%) |
| Male | 68 (87%) | 10 (13%) |
| Age (years) : | Mean: 32.56±18 (sd)<br>Median:33.09 | Mean:31.92±20.2(sd)<br>Median:30.01 |
| CT value (Median): N1 | 19.27 | 19.86 |
| N2 | 20.94 | 20.10 |

**Figure1:**
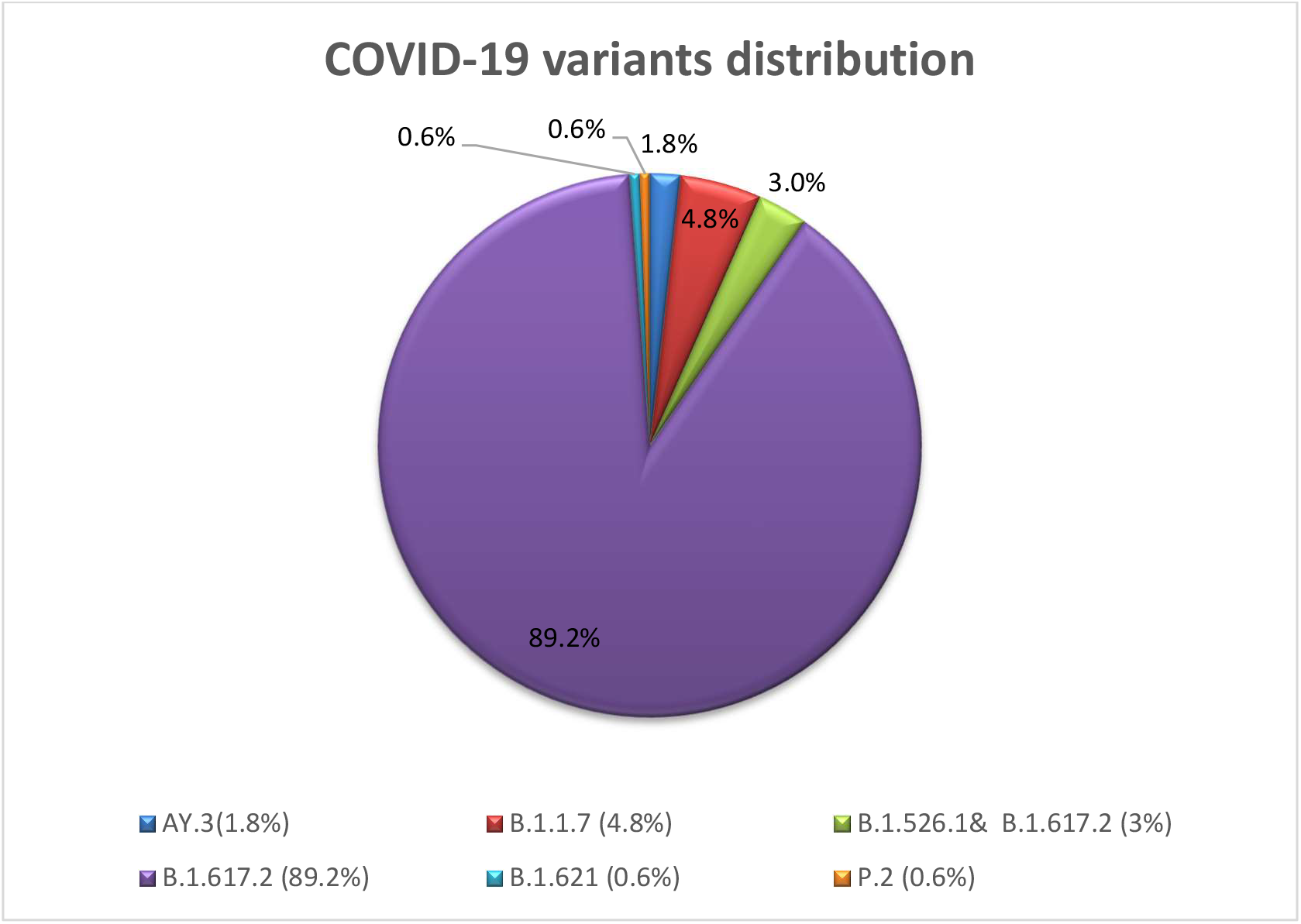
COVID-19 variant distribution

### Patient questionnaires

121 of 166 patients responded to the questionnaire to address questions of COVID-19 immunization, such as a) Were you vaccinated before testing COVID-19 positive? b) What was the date you received 1^st^ /2^nd^ dose of the vaccine? c) What kind of vaccine did you receive and from which manufacture?

38% of patients indicated that they received either first dose (4%) or full dose of vaccine (34%), and 62% of patients were not vaccinated. Among the patients who were vaccinated, 76% of patients received vaccines from Pfizer-BioNTech, 15% from Moderna, and 9% from Janssen (Figure 2). The vaccine breakthrough cases occurred in all variants with a majority of Delta variant (91%) followed by AY.3 (5%), B.1.1.7 (2%), and double variant of B.1.526.1 and B.1.617.2 (2%). Patients were diagnosed with COVID-19 positive after 4 days to 31 weeks since the first day they received the full dose of vaccine (Mean=15.15 weeks, Median=17.36 weeks, Mode=19 weeks).

**Figure 2:**
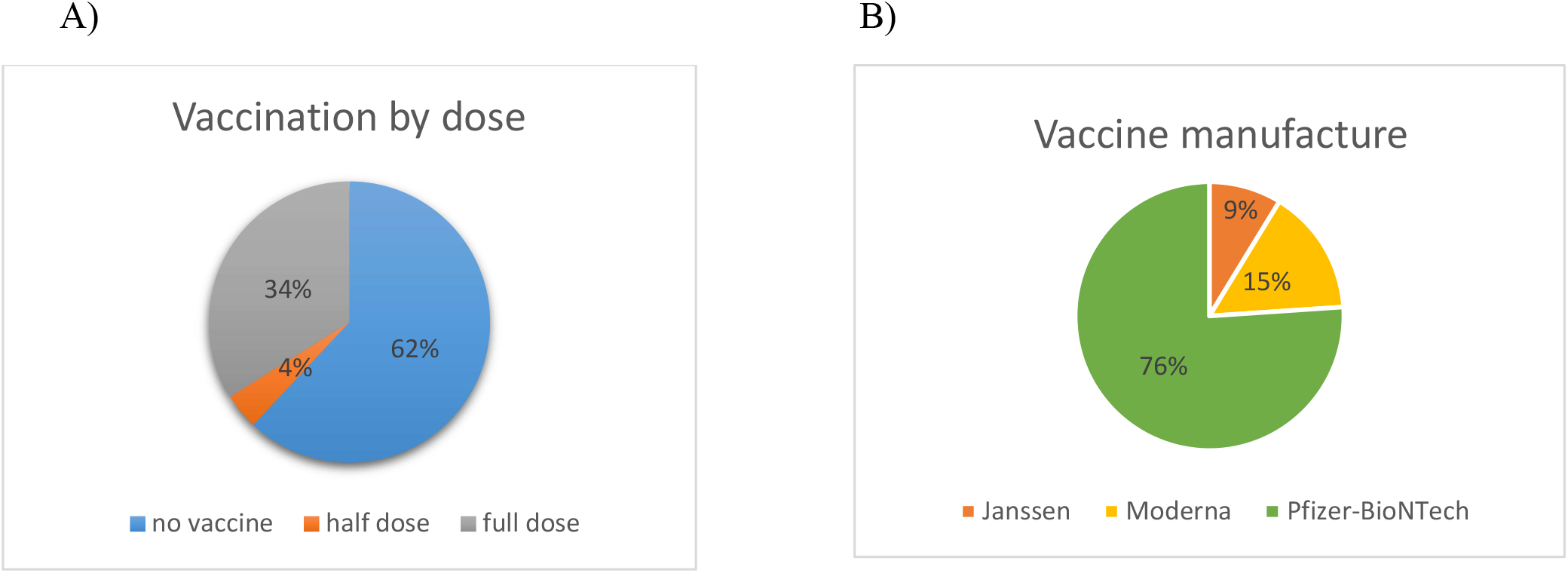
A) The pie chart represents the percentage of patients’ immunization status. B) The pie chart represents the percentage of vaccine manufactures patients received.

At the same time, 59% of patients indicated they had symptoms (e.g. fever, headache, sore throat, loss of smell/taste, fatigue, cough, shortness of breath, chest pain, etc.) when they came to our laboratory to test COVID-19, while 41% patients indicated that they came to test due to work requirements or possible exposure, but no symptoms have indicated.

### Cycle Threshold (CT) value

In general, a lower CT value reveals a higher viral load in the specimen, and a higher CT value reveals a lower viral load. As part of this study, we want to know whether the CT values are affected by any factors or provide any predictions for clinical treatment. We compared CT value between the age groups, gender, vaccinated/non-vaccinated, status of immunization, and variant groups (Delta vs other variants). A one-way ANOVA revealed that there was a significant difference in CT value between three age groups: <30 years, 31-60 years, >60 years (N1: F (2, 118) = 4.96, p=0.009; N2: F(2,118)=4.95, p= 0.009) (Figure 3). Turkey’s HSD test for multiple comparison found that the mean value of CT value was significantly different between age group <30 years and age group 31-60 years (N1: p=0.014, 95% C.I=(0.4 - 4.28);N2: p=0.012, 95% C.I=(0.5 - 4.86)). There was no statistically significant difference between the age group <30 years and age group >60 years.

**Figure 3.**
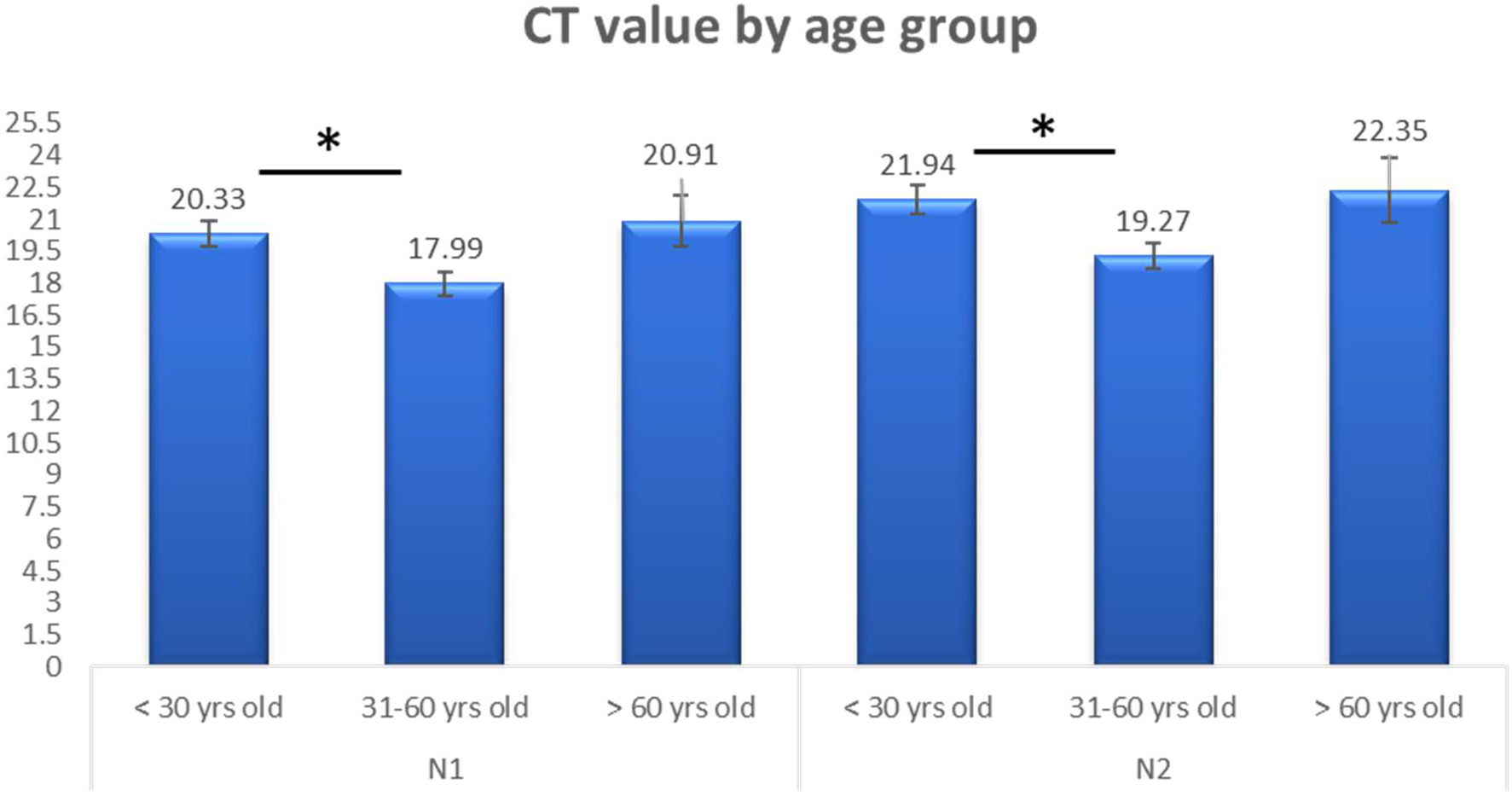
The CT value compared by the age group. A one-way ANOVA revealed that there was a significant difference in CT value between three age groups: <30 years, 31-60 years, >60 years (N1: F (2, 118) = 4.96, p=0.009; N2: F (2, 118) = 4.95, p=0.009).

Patients with symptoms shows a lower CT value than those without symptoms. The CT value was observed to be lower in the patients who received only 1^st^ dose than patients who received the full dose. The CT value was observed to be lower in the patients who received Janssen vaccine compared to the patients who received vaccines from Modera or Pfizer-BioNTech. Meanwhile, the CT value was observed to be lower for males compared for females. However, no statistically significant was identified in these observations at this point. The CT value didn’t show a significant difference between vaccine and non-vaccine groups as well.

A two-way ANOVA was performed to analyze the effect of gender and variant group (Delta and other variants) on CT value. The analysis revealed that there was a statistically significant interaction between the effect of gender and variant group (N1, F (1.117) = 3.906, p= 0.05; N2, F (1,117) = 7.402, p= 0.008) (Figure 4).

**Figure 4.**
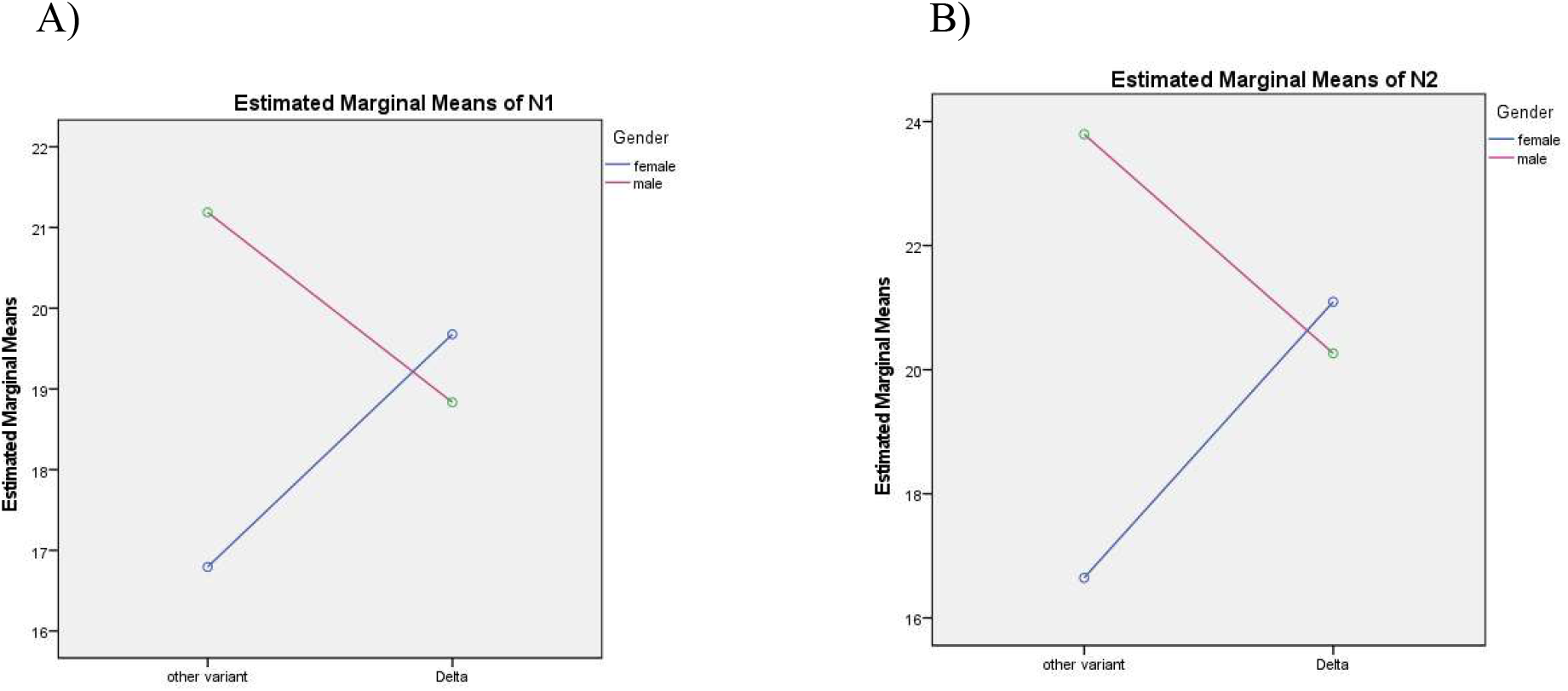
A two-way ANOVA to examine the effect of gender and variant group (Delta and other variant) on CT values. A) and B) The analysis revealed that there was a statistically significant interaction between the effect of gender and variant group (N1, F (1.117) =3.906, p=0.05; N2, F(1,117) =7.402, p=0.008).

### The association

We conducted Fisher’s exact test to check whether there is an association between the status of immunization and gender, age groups, and with/without symptoms. A significant association between vaccine and age groups was detected (p=0.000). No significant association was detected between status of immunization and gender or symptoms (Table 2).

**Table 2.** The association between the status of immunization and gender, age, and symptoms.

|  | Non vaccinated | vaccinated | P value |
| --- | --- | --- | --- |
| Gender: |  |  |  |
| Female | 33 | 26 | 0.125 |
| male | 42 | 20 |  |
| Age group: |  |  |  |
| <30 yrs old | 47 | 6 | 0.000 |
| 31-60 yrs old | 27 | 30 |  |
| >61 yrs old | 1 | 10 |  |
| Symptom: |  |  |  |
| Yes | 41 | 30 | 0.342 |
| No | 34 | 16 |  |

## Discussion

In this study, we assessed the incidence of COVID-19 variants in the communities. The result is consistent with the CDC’s prediction that the Delta variant has dominated in the communities and the country (CDC, Delta Variant, 2021).

Also, the Delta variant shows more resistance to the current vaccine regimen than other variants. The reasons may be caused by the Delta variant’s specific mutations and the nature of the vaccine. B.1.617.2 (Delta) variant is characterized by spike protein mutations T19R, Δ157-158, L452R, T478K, D614G, P681R, and D950N. Teng and Chen’s studies suggested the L452 mutation may stabilize the interaction between the spike protein and its human ACE2 receptor and thereby increase infectivity (Teng et al., 2020, Chen et al., 2020). Meanwhile, specific monoclonal antibody treatments may be less effective for treating cases of COVID-19 caused by variants with mutation of L452R (Deng et al., 2021, Corti et al., 2021).

Just before wrapping up this study, Pfizer released its data from its clinical trials. The data showed that the efficacy of its vaccine degrades by around 6% every two months after the second dose. The Kaiser Permanente Southern California research showed that the effectiveness against coronavirus infections dropped from 88% during the first months after full vaccination to 53% after ≥ 4 months and 47% after ≥ 5 months (Tratof, 2021). It is consistent with our research that patients are more susceptible to the coronavirus about 17 weeks after the first day they received their full dose of vaccine. But, the vaccine retains its potency for preventing severe illness and against hospitalization (Planas et al., 2021).

So now, Pfizer asks FDA to approve COVID booster shot and Israel is preparing for possible fourth COVID vaccine dose (Odenheimer, 2021). However, the effectiveness of the booster shot against the future new variant is unknown. Further studies are needed. Therefore, it is critical to continue to monitor the incidence of new variant and its trend. In doing so, it could help the local health unit and local authority prepare for the potential local outbreak or surge.

There are some limitations in this study. First, we were unable to examine patients’ neutralizing antibody were present after the first and second dose of vaccine, or the baseline antibody test before illness and after vaccination. Second, we were unable to conduct the physical examination, blood work, or access to patients’ medical record. We are therefore unable to obtain additional information to analyze CT values and its risk factors.

## Conclusion

This study identified the Delta variant is the dominant variant spreading in the communities. The breakthrough vaccine cases occur in a majority of Delta variant and the occurrence of the breakthrough cases was not linked to a specific vaccine manufacture. The viral load (CT value) may have an association with age. The study here supports our laboratory’s ongoing efforts to sequence the SARS-CoV-2 gene from positive patient samples to monitor the COVID-19 variant in the communities and the possible vaccine breakthrough mutations. The study also supported that the critical importance of maintaining the public health mitigation measures by wearing mask, physical distancing, daily symptom screening, regular testing, avoiding crowds and unnecessary travel, obtaining a full vaccination until herd immunity is reached at large.

## Data Availability

Data available on request from the authors.

